# Baseline Data Collection of Persons with Intellectual or Developmental Disabilities by Housing Status A Cross-Sectional Baseline Survey of Adults with Disabilities in Supported Independent and Family Living

**DOI:** 10.64898/2026.05.11.26352919

**Authors:** Sean D. Cleary

**Affiliations:** Department of Epidemiology, George Washington University, Washington, DC

## Abstract

This report presents findings from a baseline cross-sectional survey of autistic adults and adults with intellectual and/or developmental disabilities connected to Our Stomping Ground (OSG), a nonprofit organization in northern Virginia committed to supported independent living. The survey was administered in 2022–2023 as the first wave of a planned longitudinal study, with the primary goal of establishing a comparable starting point between two groups: adults who were living — or preparing to live — independently in an OSG apartment building, and adults with disabilities who were continuing to live at home with family.

A total of 76 adults completed the survey out of 98 potential participants. The two groups were well-matched at baseline across a wide range of characteristics, which is exactly what a sound longitudinal design requires. This comparability means that when follow-up data are collected, any observed differences between the groups can be more confidently attributed to the experience of independent living rather than to pre-existing differences.

The main findings at baseline were as follows:

**Key Baseline Findings:**

- The two groups — apartment residents and those living at home — did not differ significantly on any demographic, health, or psychosocial characteristic, with just two exceptions: more apartment residents completed the survey independently (93% vs. 69%), and, as expected, the groups differed on who they currently lived with.
- The most common diagnoses were autism spectrum disorder (69%), developmental disorders (82%), and intellectual or cognitive disorders (49%). The mean age of participants was 28 years.
- Mental health counseling was already the most common current treatment (41%), consistent with its documented importance to this population.
- Physical activity and BMI profiles were similar across groups: 24% low, 40% moderate, and 37% high physical activity; 50% normal weight, 24% obese.
- Quality of life, as measured by the WHO Quality of Life–BREF, was broadly positive: 84% rated their overall quality of life as good or very good.
- Social support, self-esteem, coping, loneliness, and community contribution were all similar across the two groups, providing a solid, equitable baseline for future comparison.
- At least one in four participants reported needing professional support in most areas of daily life, including employment, finances, transportation, and counseling — underscoring the level of support that makes independent living achievable.

These findings provide the empirical foundation for the longitudinal phase of this study and offer a meaningful snapshot of the health, well-being, and support needs of autistic adults and adults with INTELLECTUAL AND/OR DEVELOPMENTAL DISABILITIES at a pivotal life stage. They confirm that OSG is serving a population with real and multifaceted needs — and that both groups enter the study on genuinely comparable footing.

## I. Background and Context

Adults with disabilities face unique challenges in achieving independence and maintaining their well-being. The movement towards independent living, versus living with their families, presents different support needs as well as potential benefits. It is therefore important to study adults with disabilities living independently and those continuing to live at home with their families. Understanding the experiences of adults with disabilities in these living arrangements can inform policies and interventions aimed at enhancing their autonomy and quality of life.

## II. Study Objectives

The purpose of the baseline survey was to provide a starting point to assess and monitor changes in health, mental health, and quality of life over time. Baseline data collected from adults with disabilities living independently and living at home offers a comparative benchmark for evaluating the impact of interventions or support services. Studying baseline characteristics can identify factors that contribute to positive outcomes and inform targeted interventions to address specific needs. Survey results provide a foundation for future research by establishing a baseline against which longitudinal studies can measure changes and evaluate effectiveness. In addition, understanding baseline levels can facilitate resource allocation and planning of support systems for individuals with disabilities living independently or with their families.

More specifically, we explored the prevalence of chronic conditions, disabilities, and functional limitations to get a picture of physical well-being and identify any specific health concerns. Mental health conditions such as depression, anxiety, and stress were assessed to understand the psychological well-being and emotional functioning of participants. We also assessed baseline quality of life to evaluate participants’ overall satisfaction with life, social relationships, and community participation. Other characteristics were assessed as well, including demographic factors such as age, gender, and education level, social support networks, and access to support services that may influence health, mental health, and quality of life outcomes.

The rationale for the domains selected is as follows: understanding the baseline health status of adults with disabilities is crucial for identifying specific needs and developing targeted interventions; mental health is a critical aspect of overall well-being, yet there is limited research on the mental health status of adults with disabilities in different living arrangements; assessing baseline quality of life can provide insights into subjective well-being and help identify areas for improvement; and assessing demographic factors, social support networks, and other assets can help contextualize health outcomes and inform the development of personalized support systems.

## III. Methodology

### A. Study Design

A cross-sectional survey design allows for the collection of data at a single point in time, providing a snapshot of the baseline characteristics of adults with disabilities by housing status. This design is suitable for examining baseline levels of health, mental health, quality of life, and other characteristics, as it allows for a comprehensive assessment of these variables within the study population.

### B. Participants

Potential participants included adults (18 years and above) with disabilities. Disabilities encompass a range of physical, sensory, cognitive, and developmental impairments that may impact daily functioning and require support. Among those with disabilities, we sought a group that currently was, or in a short period of time would be, living independently in one of OSG’s apartment buildings. A comparison group included adults with disabilities who would continue to live at home. All potential participants were identified through OSG staff.

Although this was a convenience sample, the recruitment process involved multiple strategies to ensure a diverse and representative sample. Interested participants were sent a flyer and instructed to contact a representative from George Washington University (GW) for more information. Potential participants were then emailed a unique ID number and a link to complete the survey after providing informed consent. All procedures conformed to ethical standards for participant confidentiality and research integrity, as approved by GW IRB (#NCR224387).

The original target sample size was 100, with 50 participants in each group, assuming a medium to large effect size (above 0.6), power greater than 80%, and alpha at 0.05.

### C. Data Collection

A structured questionnaire was developed and pretested by George Washington University staff. Study data were collected and managed using REDCap electronic data capture tools hosted at The George Washington University. Each potential participant was provided a unique ID and a link to the informed consent form and survey. Surveys could be self-administered or completed with assistance from a parent or OSG staff. When possible, standardized assessments were selected to obtain objective, validated measures across health and psychosocial domains.

### D. Data Analysis

The collected data were cleaned, coded, and entered into SAS statistical software for analysis. Descriptive statistics — including means, frequencies, and percentages — were used to provide a comprehensive overview of baseline characteristics. Inferential statistical tests, including chi-square and ANOVA tests, were used to examine associations and differences between groups. Consistent with the study design, non-significant differences between the two groups at baseline were expected. Once longitudinal follow-up data are collected, inferential statistics will be used to evaluate the impact of the independent living intervention after adjusting for baseline differences. Significant findings are highlighted in the text below.

## IV. Results

Overall, 76 out of 98 potential participants completed the survey. Of the 47 participants living at home, 32 (68%) completed all or part of the survey. Of the 51 participants living in apartments, 44 (86%) completed a survey. The higher response rate among apartment residents may be partially explained by anecdotal reports from OSG staff that participants taking the survey enjoyed the process and the time they had to talk about their lives and living experiences. There was no monetary incentive to complete the survey.

### A. Demographics

Preliminary demographic statistics for the entire sample are described below. Table 1 shows the demographic variables by housing status. To maintain confidentiality, response options with fewer than 5 respondents are masked and indicated by (**).

**Table 1.** Baseline Demographics — Overall and by Housing Status.

| Characteristic | Overall (N=76) | Family Home (N=32) | Apartment (N=44) | Significant Differences |
| --- | --- | --- | --- | --- |
| Age (years), mean (SD) | 28.0 (9.3) | 25.5 (6.0) | 30.0 (10.9) | n.s. |
| Survey completed independently, % (n) | 82.7% (62) | 68.8% (22) | 93.0% (40) | **p < 0.01 ✓** |
| Sex: Male, % (n) | 53.3% (40) | 59.4% (19) | 48.8% (21) | n.s. |
| Race/Ethnicity, % (n) |  |  |  |  |
| White non-Hispanic | 70.3% (52) | 68.8% (22) | 71.4% (30) | n.s. |
| Black non-Hispanic | 12.2% (9) | ** | 16.7% (7) |  |
| Asian non-Hispanic | 9.5% (7) | ** | ** |  |
| Other | 8.1% (6) | ** | ** |  |
| **Disability Diagnoses, % (n)** |  |  |  |  |
| Developmental Disorder | 54.1% (40) | 87.1% (27) | 59.5% (25) | n.s. |
| Autism Spectrum Disorder | 68.5% (50) | 71.9% (23) | 65.9% (27) | n.s. |
| Intellectual/Cognitive Disorder | 49.3% (35) | 43.3% (13) | 53.7% (22) | n.s. |
| Language: Uses longer sentences, % (n) | 83.8% (62) | 87.5% (28) | 81.0% (34) | n.s. |
| **Currently lives with parent/relative, % (n)** | 36.8% (28) | 87.5% (28) | ** | **p < 0.0001 ✓** |
| **Education Level, % (n)** |  |  |  |  |
| Less than high school | 13.5% (10) | ** | ** | n.s. |
| High school diploma / GED | 48.6% (36) | 50.0% (16) | 47.6% (20) |  |
| Some college | 19.3% (15) | 18.8% (6) | 21.4% (9) |  |
| Bachelor's degree | 8.1% (6) | ** | ** |  |
| **Employment, % (n)** |  |  |  |  |
| Part-time with support | 20.1% (15) | ** | 26.2% (11) | n.s. |
| Part-time (no support) | 24.3% (18) | 23.5% (8) | 25.0% (8) |  |
| Unemployed | 25.7% (18) | 18.9% (7) | 28.6% (12) |  |
| **Health Insurance, % (n)** |  |  |  |  |
| Medicaid | 31.4% (22) | 27.6% (8) | 34.2% (14) | n.s. |
| Medicare | 12.9% (9) | ** | 14.6% (6) |  |
| Parent's plan | 41.4% (29) | 44.8% (13) | 39.0% (16) |  |
| Other | 14.2% (10) | ** | ** |  |

Note. n.s. = not statistically significant at $p < 0.05$ ; \*\* = masked due to fewer than 5 participants per cell. SD = standard deviation. ✓ indicates statistically significant difference.

**Table 2.** Number of Medical Conditions, Past and Current Treatments.

| Variable | N | Missing | Mean | Std Dev | Minimum | Maximum |
| --- | --- | --- | --- | --- | --- | --- |
| Psychological conditions | 76 | 0 | 1.26 | 1.35 | 0 | 6 |
| Developmental conditions | 76 | 0 | 1.33 | 1.20 | 0 | 5 |
| Physical health conditions | 76 | 0 | 1.55 | 1.46 | 0 | 6 |
| Past treatments | 76 | 0 | 5.54 | 2.70 | 0 | 10 |
| Current treatments | 76 | 0 | 1.46 | 1.42 | 0 | 7 |

Note. No statistically significant differences by housing status were observed. SD = standard deviation.

The mean age of respondents was 28 years. Overall, 53% were male, 44% female, and 3% identified as transgender. In terms of race and ethnicity, 70% were white non-Hispanic, 12% Black non-Hispanic, 9.5% Asian/Pacific Islander non-Hispanic, 5% Hispanic, and 3% other. Fifty-four percent were legally independent, with a parent or sibling serving as legal guardian for the remainder. With respect to diagnoses, 82% reported a developmental disorder, 69% an autism spectrum disorder, and 49% an intellectual or cognitive disorder. The majority (84%) of respondents reported full language fluency. Eighty-three percent of respondents completed the survey on their own. Virtually all respondents (99%) had never been married, and all reported having health insurance coverage.

Only two demographic characteristics differed significantly by housing status: the role of the respondent in completing the survey (p < 0.01) — significantly more apartment residents completed it independently — and who respondents currently lived with (p < 0.0001), as expected. **There were no statistically significant differences on any other demographic characteristic by housing status**.

### B. Medical Conditions and Treatment Experiences

Respondents were asked to identify physical health (n=6 items), mental health (n=6 items), and developmental (n=5 items) conditions they were currently experiencing, as well as past and current treatments. Positive responses were summed within each category. There were no statistically significant differences by housing status in the prevalence or number of self-reported medical conditions or treatment experiences.

The most commonly reported conditions were a learning disability (53%), ADHD (47%), allergies (33%), and language delays (24%). Approximately one in five respondents reported depression, OCD, social anxiety, motor delays, speech delays, and sleep disorders. In terms of past treatments, 79% reported speech and language therapy, 71% occupational therapy for fine motor skills, 67% mental health counseling, 61% occupational therapy, and 61% social skills groups. Among current treatments, the most common were mental health counseling (41%), social skills groups (33%), and biomedical treatments (28%).

### C. Health Services Utilization

Respondents were asked to report the number of health service visits during the past year. Table 3 includes the mean number of visits for each service type. There were no statistically significant differences by housing status in any category of health service utilization.

**Table 3.** Health Service Utilization — Mean Number of Visits During the Past Year.

| Service Type | N | Missing | Mean | Std Dev | Minimum | Maximum |
| --- | --- | --- | --- | --- | --- | --- |
| Hospital Emergency Room visits | 71 | 5 | 0.48 | 1.39 | 0 | 10 |
| Doctor visits | 70 | 6 | 5.87 | 11.42 | 0 | 70 |
| Mental health professional visits | 70 | 6 | 6.70 | 13.17 | 0 | 79 |
| Specialist visits | 70 | 6 | 3.31 | 9.58 | 0 | 70 |
| Vision testing | 71 | 5 | 0.80 | 1.44 | 0 | 11 |
| Dental visits | 71 | 5 | 2.27 | 2.32 | 0 | 12 |

Note. No statistically significant differences by housing status were observed. SD = standard deviation.

**Table 4.** Mean Social Support Score by Domain and Social Source.

| Domain / Source | N | Mean | Std Dev | Minimum | Maximum | Cronbach's Alpha |
| --- | --- | --- | --- | --- | --- | --- |
| **By Domain** |  |  |  |  |  |  |
| Talk | 71 | 4.03 | 0.75 | 1 | 5 | 0.75 |
| Listen | 71 | 4.04 | 0.70 | 2 | 5 | 0.70 |
| Understand | 71 | 4.03 | 0.63 | 2 | 5 | 0.63 |
| Feel better | 71 | 4.00 | 0.73 | 2 | 5 | 0.74 |
| Suggestions | 71 | 3.95 | 0.68 | 2 | 5 | 0.70 |
| Money | 71 | 3.40 | 0.82 | 2 | 5 | 0.74 |
| School work | 70 | 3.61 | 0.78 | 1 | 5 | 0.75 |
| Travel | 71 | 3.73 | 0.74 | 2 | 5 | 0.69 |
| Health | 71 | 3.66 | 0.71 | 2 | 5 | 0.63 |
| Friends | 71 | 3.75 | 0.72 | 2 | 5 | 0.67 |
| **By Social Source** |  |  |  |  |  |  |
| Parent | 71 | 4.52 | 0.51 | 3 | 5 | 0.88 |
| Friend | 71 | 3.69 | 0.81 | 1 | 5 | 0.93 |
| School / work adults | 71 | 3.57 | 0.73 | 2 | 5 | 0.89 |
| Community adults | 71 | 3.49 | 0.94 | 1 | 5 | 0.95 |
| **Total score (all 40 items)** | **71** | **3.82** | **0.59** | **2** | **5** | **0.95** |

Note. Scale rated 1 (strongly disagree) to 5 (strongly agree). No statistically significant differences by housing status were observed. SD = standard deviation.

**Table 5.** WHO Quality of Life Scales — Baseline Scores.

| Domain | N | Missing | Mean | Std Dev | Minimum | Maximum | Range |
| --- | --- | --- | --- | --- | --- | --- | --- |
| Base score (overall QoL) | 71 | 5 | 74.65 | 19.25 | 25 | 100 | 75 |
| Physical Health | 71 | 5 | 69.92 | 15.42 | 36 | 100 | 64 |
| Psychological Health | 71 | 5 | 65.14 | 16.89 | 21 | 100 | 79 |
| Social Relationships | 70 | 6 | 52.62 | 26.03 | 0 | 100 | 100 |
| Environment | 71 | 5 | 73.42 | 16.31 | 31 | 100 | 69 |

Note. Higher scores indicate greater quality of life. No statistically significant differences by housing status were observed. SD = standard deviation.

**Table 6.** WHO Disability Assessment Schedule 2.0 (WHODAS 2.0) — Baseline Scores.

| Domain | N | Missing | Mean | Std Dev | Minimum | Maximum | Range |
| --- | --- | --- | --- | --- | --- | --- | --- |
| Cognition | 68 | 8 | 43.01 | 23.34 | 0 | 85 | 85 |
| Mobility | 68 | 8 | 20.68 | 27.02 | 0 | 125 | 125 |
| Self-care | 68 | 8 | 23.97 | 35.75 | 0 | 140 | 140 |
| Getting along with others | 67 | 9 | 54.98 | 32.86 | 0 | 125 | 125 |
| Life activities (household) | 68 | 8 | 57.50 | 43.03 | 0 | 160 | 160 |
| Community participation | 68 | 8 | 45.77 | 27.76 | 0 | 100 | 100 |
| **Simple total score** | **68** | **8** | **75.48** | **22.44** | **35** | **140** | **105** |
| **Normed total score (range 0–100)** | **68** | **8** | **40.73** | **22.08** | **0** | **100** | **100** |

Note. Higher scores indicate greater level of disability. No statistically significant differences in subscale or total scores by housing status were observed. SD = standard deviation.

### D. Health Care Not Received

There were too few responses to the health services not received items during the past year to report meaningfully. Future data collection should consider alternative approaches to better capture unmet health care needs in this population.

### E. Physical Activity and Obesity Levels

The International Physical Activity Questionnaire (IPAQ, short form) was used to assess type, frequency, and duration of physical activity, and to classify respondents into low, moderate, and high categories. There were no statistically significant differences in physical activity level by housing status. Overall, 24% of participants reported a low level, 40% a moderate level, and 37% a high level of physical activity.

Self-reported height and weight were used to calculate body mass index (BMI), classified into four categories based on CDC guidelines. While there were no statistically significant differences by housing status in BMI level, overall 9% were classified as underweight, 50% normal weight, 17% overweight, and 24% obese at baseline.

### F. Support for Daily Living Activities

Respondents were asked whether they currently received any professional support in nine areas of daily life. Overall positive responses were as follows: 38% daily living activities, 36% counseling/therapy, 34% community and transportation, 32% employment, 30% social support, 26% health care, 26% finances, 20% for finding housing, and 18% for life coaching. Overall, 25% or more of adults with disabilities reported needing professional assistance in most areas of daily living.

There were four statistically significant differences in the need for professional support by housing status. Apartment residents were significantly more likely to receive support for counseling/therapy (46% vs. 22%; p < 0.05), health care (36% vs. 13%; p < 0.05), personal finances (36% vs. 13%; p < 0.05), and housing (30% vs. 6%; p < 0.05). These differences reflect the real demands of managing one’s own household and health independently, and the legitimate role of formal supports in making that independence sustainable.

### G. Social Support Across Domains

Social support is critical for successful transition into adulthood. Respondents rated on a Likert-type scale (strongly disagree to strongly agree) whether they received support across 10 functional areas (talking, listening, understanding, emotional support, suggestions, money, school/work, travel, health, and friends) from four social sources (parents, friends, school/work adults, and community adults). Mean scores were calculated for each area and source, as well as a total score across all 40 items. The total score demonstrated excellent internal consistency reliability (Cronbach’s alpha = 0.95). There were no statistically significant differences in social support by housing status.

### H. WHO Quality of Life Scales

The WHO Quality of Life–BREF (WHOQOL-BREF) was used to measure quality of life across four domains: physical health, psychological health, social relationships, and environment. Scores were normed from 0 to 100 (higher = better quality of life). Overall, 84% of respondents rated their quality of life as good or very good, and 67% reported being satisfied or very satisfied with their health. There were no statistically significant differences in any quality of life score by housing status.

### I. WHO Disability Assessment Schedule 2.0

The WHO Disability Assessment Schedule 2.0 (WHODAS 2.0, 32-item short form) assessed functioning across six domains: cognition, mobility, self-care, getting along with others, life activities, and community participation. Higher scores indicate greater functional limitations. There were no statistically significant differences in WHODAS 2.0 subscale scores or the normed total score by housing status.

The one statistically significant finding in this section was in the disability days items: participants living at home reported significantly more days of total inability to carry out usual activities in the past 30 days (mean 3.7 days, SD 7.2) than apartment residents (mean 1.1 days, SD 2.3; p < 0.05). This difference warrants monitoring in the longitudinal phase of the study.

### J. Other Measures: Free Time Activities, Self-Esteem, Coping, Contribution, and Loneliness

#### J.1. Free Time Activities

Respondents were asked how they spent their free time. Overall, 86% reported spending time with family, 66% with friends, 58% attending disability groups, 42% in online communities, 38% in classes, and 25% volunteering. There were no statistically significant differences in free time activities by housing status.

#### J.2. Self-Esteem

The 10-item Rosenberg Self-Esteem Scale was used. Summary scores were calculated by reverse-scoring five items and summing all ten. The mean score was 36.9 (SD = 7.7, range 14 to 50). Cronbach’s alpha was 0.87, indicating high reliability. There was no significant difference in self-esteem by housing status.

#### J.3. Coping

A 13-item scale assessed both positive (active) and negative (avoidant) coping behaviors. The mean score for positive coping was 20.5 (SD = 6.7, range 9 to 36) and for negative coping was 3.8 (SD = 3.0, range 0 to 16). Cronbach’s alpha was 0.89 for positive coping and 0.80 for negative coping, indicating strong reliability for both. There was no significant difference in positive or negative coping by housing status.

#### J.4. Contribution

A 14-item scale measuring contribution to one’s family, peers, school, church, and community was included. Contribution is considered a positive developmental asset according to Lerner’s Positive Youth Development framework. All items were summed for a total score of mean 10.8 (SD = 7.9, range 0 to 45; alpha = 0.76).

There was no significant difference in contribution by housing status.

#### J.5. Loneliness

The 16-item UCLA Revised Loneliness Scale was used. After reverse-scoring seven items, all 16 responses were summed for a total loneliness score (mean = 31.5, SD = 9.7, range 16 to 52; alpha = 0.90). There was no statistically significant difference in loneliness by housing status.

## V. Discussion

### A. Primary Findings

The purpose of this study was to collect baseline levels of health, mental health, quality of life, and other characteristics among adults with disabilities living independently or living with their families. The main finding is straightforward: with the exception of the role of respondent completing the survey and who respondents currently lived with, **there were no statistically significant differences between the two groups at baseline**. This is exactly the outcome a well-designed longitudinal study requires. It means that once follow-up data are collected, changes in outcomes across the two groups can be more directly attributed to the experience of independent living rather than to pre-existing differences between participants.

Notable baseline characteristics are discussed by domain in the Results section above. Taken together, the data paint a picture of a population with genuine needs across multiple areas of daily life — health, mental health, employment, finances, and social connection — and with meaningful strengths in areas such as social support, quality of life, and community engagement. Both groups enter the longitudinal phase of the study on equivalent footing, which positions the follow-up data to answer the questions that matter most about the benefits of supported independent living.

### B. Limitations

Several limitations should be considered when interpreting these findings. The cross-sectional design provides only a snapshot of baseline characteristics. The convenience sample, restricted to one geographic area and a single organization, may limit the generalizability of findings to the broader population of autistic adults or adults with INTELLECTUAL AND/OR DEVELOPMENTAL DISABILITIES. The limited sample size may reduce statistical power, particularly for subgroup analyses. Reliance on self-report data introduces the potential for social desirability bias or subjective interpretation. Despite these limitations, this study provides valuable insights into the baseline well-being and support needs of this population, and establishes a rigorous foundation for the longitudinal follow-up.

### C. Future Directions

Conducting longitudinal research to examine changes over time in health, mental health, quality of life, and social support will provide a deeper understanding of the trajectories and dynamics of well-being among adults with disabilities. Supplementing quantitative findings with qualitative or mixed-methods research can offer a more nuanced understanding of participants’ lived experiences and the factors influencing their outcomes. Designing and evaluating targeted interventions — particularly those addressing mental health access, employment, and financial support — will contribute to evidence-based practice and inform policy decisions for this population.

This study makes a meaningful contribution to the literature by providing a comprehensive, empirically grounded examination of the baseline characteristics of autistic adults and adults with INTELLECTUAL AND/OR DEVELOPMENTAL DISABILITIES in community-based supported independent living settings. It expands the evidence base for programming, policy, and longitudinal research, and establishes the foundation for understanding what changes — and what drives those changes — over time.

## Data Availability

All data produced in the present work are contained in the manuscript

